# CLINICAL TRIAL FOR LEVOFLOXACIN VERSUS AMOXYCILLIN/CLAVULANIC ACID COMBINED WITH AZITHROMYCIN IN TREATMENT OF COMMUNITY ACQUIRED PNEUMONIA

**DOI:** 10.1101/2023.07.14.23292686

**Authors:** Vincent Musungu, Daniel Onguru, Patrick Onyango

## Abstract

**Background:** Community acquired pneumonia (CAP) is an important cause of mortality and morbidity worldwide. Early initiation of antibiotics is highly recommended. In most CAP cases, multiple drug options are increasingly becoming available, but there is often a lack of evidence that allows for direct comparison of the efficacy of one drug versus another. The purpose of this study was to investigate the effectiveness of antibiotics so as to rationalize outpatient treatment of community acquired pneumonia among patients with previous antibiotic exposure.

**Aim:** The main objective was to compare treatment outcomes using oral levofloxacin alone and combined azithromycin and amoxicillin/Clavulanic acid in outpatient treatment of Community acquired pneumonia.

**Methods:** This study was a prospective longitudinal design. Patients diagnosed to have CAP were put into first and second treatment groups. Community acquired pneumonia was diagnosed according to America Thoracic Society criteria. Sample size of 76 was arrived at by Cochran’s formular. Variation in white blood cell counts at baseline and after treatment in the two treatment groups was analysed using ANOVA whereas differences in treatment outcome was analysed using the independent t-test.

**Results:** The findings of this study suggest that combination of azithromycin and amoxycillin/clavulanic acid was associated with statistically significant faster resolution of chest pains and cough (mean 1.7 and 3.14 days respectively) compared to levofloxacin group (mean 2.21 and 3.71 days respectively) in patients who had community acquired pneumonia (p=0.009). However, in the two treatment groups, there was no difference in the meantime to fever resolution, time to crackles subsidence, resolution of difficulty in breathing and change in white blood cell count.

**Conclusions:** Based on the quantitative analysis and methodology used by the current study, it can be concluded that although both treatment groups demonstrated similar effectiveness, combination of amoxicillin/clavulanic acid and azithromycin showed a modest superiority in relation to rate of symptom resolution and change in white blood cell count.

## Introduction

Both community- and hospital-acquired pneumonia is an inflammation of the lung parenchyma (Jain et al., 2022). The term “ community-acquired pneumonia” (CAP) refers to an acute illness that develops within two weeks, does not require hospitalization, and is clinically accompanied by at least two respiratory symptoms, such as a productive cough, dyspnea, chest pain, hemoptysis, or fever, as well as consolidation on a chest x-ray (Muthumbi et al., 2017).

Adults who present with suspected CAP get empirical antimicrobial chemotherapy in accordance with the pertinent national recommendations, with age, comorbidities, and the severity of the disease serving as the main determinants of the antibiotic class and method of administration (Hoque et al., 2013). For previously healthy patients who haven’t taken any antibiotics in the three months before presentation, the American Thoracic Society (ATS) and the Infectious Diseases Society of America (IDSA) both advocate monotherapy with macrolides or doxycycline in outpatient settings. For individuals with CAP and a recent antibiotic use, a combination of an antipneumococcal -lactam like amoxycillin and a macrolide like azithromycin or a respiratory fluoroquinolone like levofloxacin is advised (Lee et al., 2018; Mandell et al., 2007).

Antibiotic resistance is a risk factor for recent antibiotic usage within the previous three months, and if the patient has recently been exposed to the same antibiotic, it is not advised for continuous or recurrent use or of the same class of broad spectrum. There are a growing number of pharmacological alternatives, but frequently there isn’t enough direct clinical trial data to compare the efficacy of different drugs. However, decision-making in clinical practice requires knowledge of the relative efficacy between different drugs (Kim et al., 2014; Mandell et al., 2007). Given the challenge posed by multiple drugs available in treatment of CAP, this study compared the effectiveness oral levofloxacin when used alone and amoxicillin/clavulanic acid combined with azithromycin in the treatment of CAP in the outpatient for those patients who have been exposed to antibiotics within the last three months.

## Materials and methods

### Study Design

The study utilized prospective longitudinal study design. Longitudinal designs involve repeated observation of the same participants to follow change over time. In the current study patients who were diagnosed with community acquired pneumonia were allocated to one of the usual treatment groups and observed for five days at an interval (Figure 1). The study was carried out between march 6, 2022 and December 18, 2022 at St Monica hospital Kisumu, Kenya.

**Figure 1:**
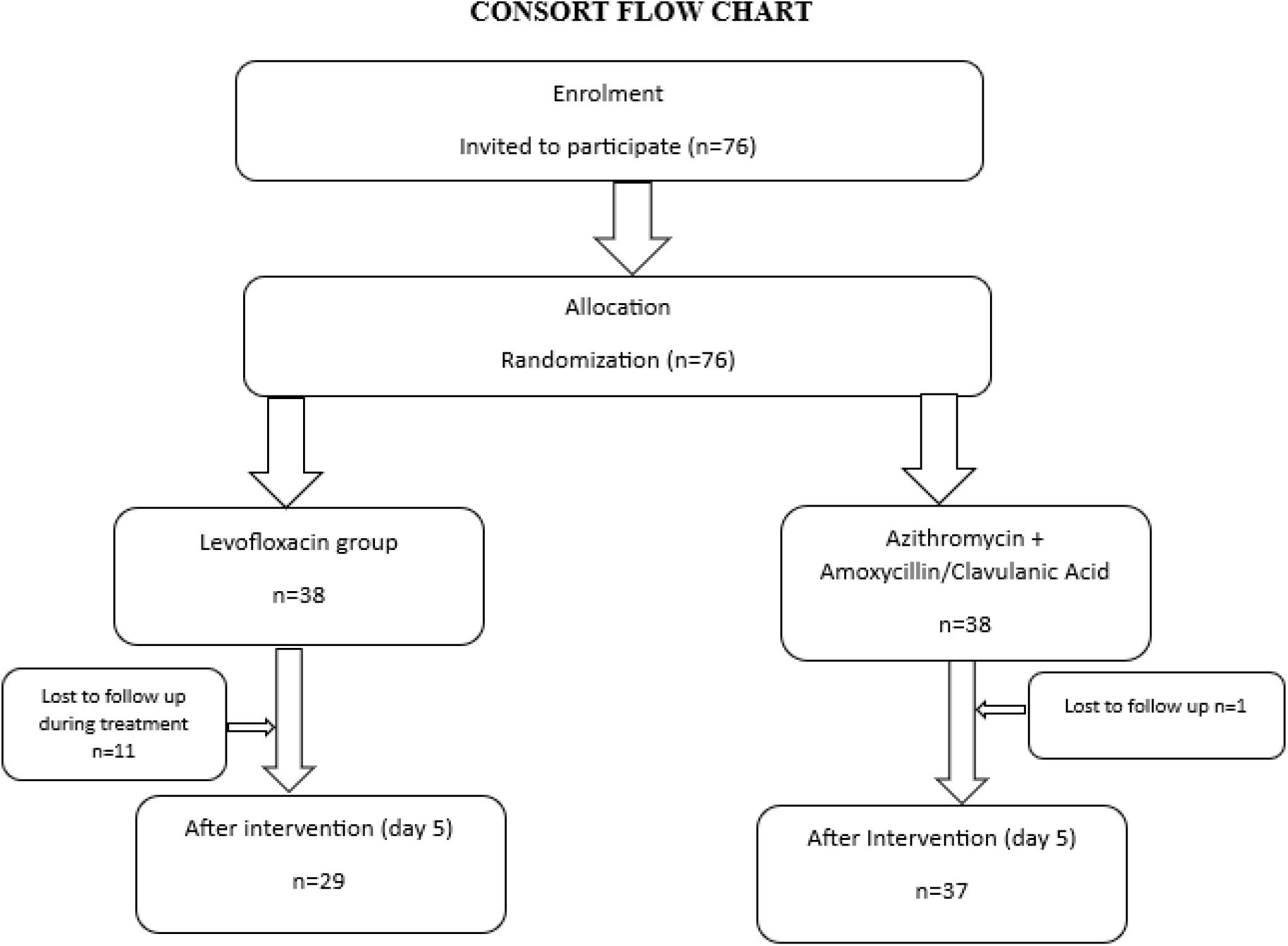
Consort flow chart.

This design was chosen to allow for a comparison of the efficacy of oral levofloxacin alone versus oral azithromycin and amoxicillin/clavulanic acid in the treatment of community-acquired pneumonia in patients who were observed at intervals during treatment. These drugs are already in use, and there is no new drug involved. On days 1, 3, and 5, the patients’ information on clinical parameters was repeatedly gathered. The change in clinical parameters of patients during the observation was used to determine effectiveness. The clinical parameters in the study included fever, cough, chest pain, shortness of breath, physical findings of crackles, and white blood cell count. A drug was considered effective if taking it resulted in the resolution of clinical parameters at the end of the treatment period. Participants were recruited as they came to the hospital for outpatient care between March 2022 and December 2022 provided, they consented and met the eligibility criteria.

The study population consisted of patients diagnosed to have community acquired pneumonia who were put in levofloxacin group and azithromycin and amoxycillin/clavulanic acid group. Community acquired pneumonia was diagnosed according to America Thoracic Society (ATS) criteria in which signs and symptoms of pneumonia included at least two of the following:

a. fever (axillary temperature > 37.5°C),
b. cough for less than 14 days,
c. chest pain,
d. shortness of breath,
e. physical findings of consolidation,
f. white blood cell count >15000/ul or <5000/ul,
g. Chest x-ray showing evidence of lung infection (pulmonary opacity).

### Sampling process

Every patient diagnosed and treated in outpatient department who gave a written consent to participate was enrolled in the study and randomly assigned to one of the treatment groups. Minors below 12 years were excluded from the study. The researcher with the help of research assistants enrolled every eligible participant as they came until 76 participants were enrolled in the study **(Figure 2)**. Patients were randomly assigned into each treatment group to get 38 patients in each group.

**Figure 2:**
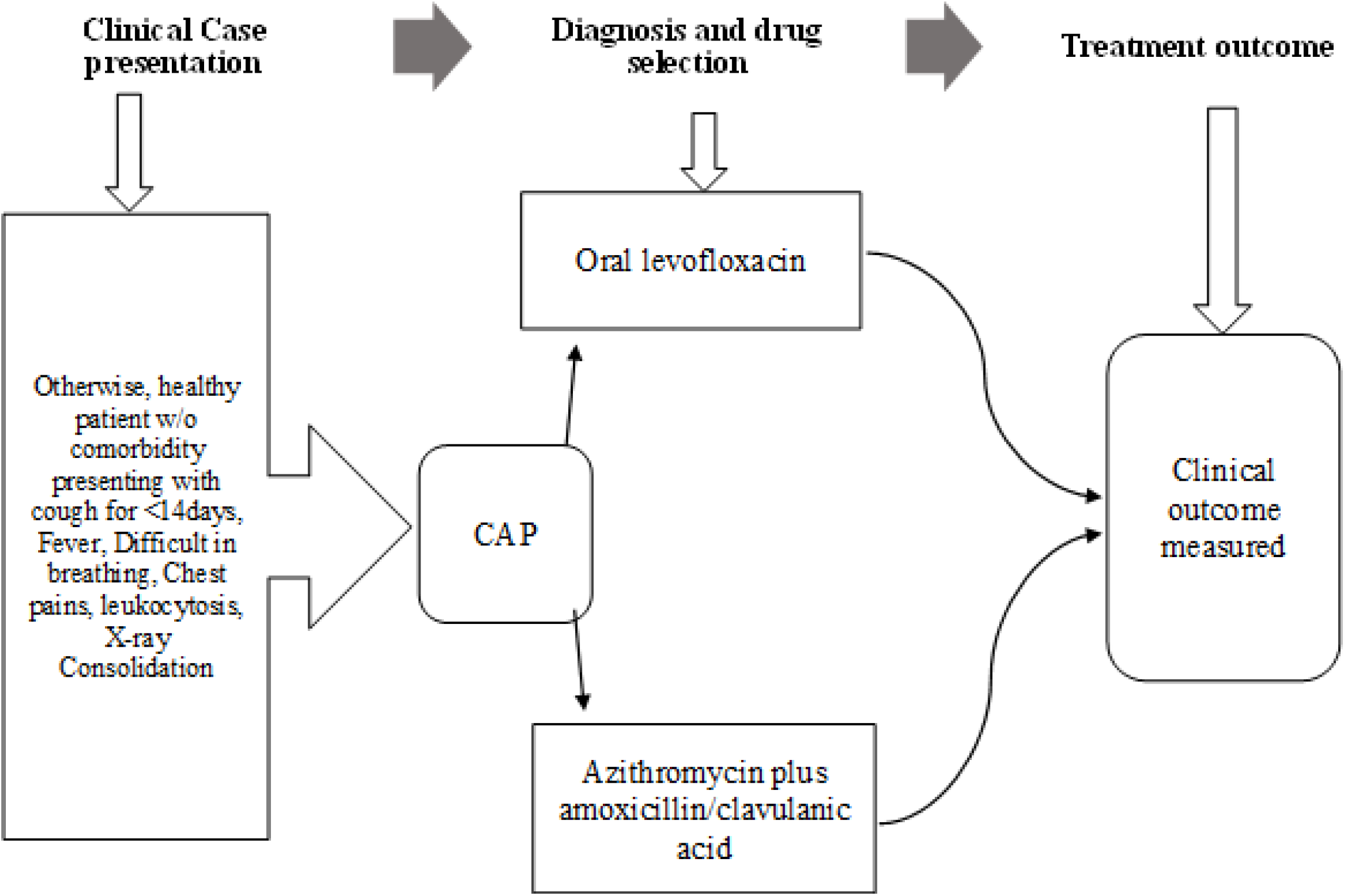
Sampling process.

### Data analysis

Variation in white blood cell counts at baseline and after treatment in the two treatment groups was done by ANOVA, Analysis of treatment outcome variation in the second treatment group (azithromycin amoxicillin/CA) was done by independent t-test. A p-value of <0.05 was considered as significant. Data was analysed using SPSS Version 26.

### Ethical consideration

This study was licensed by national commission for science, innovation and technology vide license number NACOSTI/P/22/15077. Jaramogi Oginga Odinga University of science and technology Board of Postgraduate Studies, approved this study vide approval letter reference number: 152/4071/2017. This study was approved by ethical committee reference number B0734432021. Each patient was explained to about the study including benefits and risks. Those who accepted were given the consent form to sign written informed consent form. Minors were excluded from the study. All information obtained from the study was kept in safe box by the researcher. No patient identifiers were collected during and after the study. The researcher provided adverse events notification form in case a patient experienced allergies or reaction to the drugs in the study. There was no adverse event documented at the end of the study.

## RESULTS

### Demographic characteristics of respondents

The patients were categorized into two treatment groups i.e., oral levofloxacin-based therapy, 29(43.9%) and dual Azithromycin and Amoxicillin/Clavulanic acid-based therapy, 37(56.1%) and compared during the study. About fifty percent, 33(50%) of the patients were aged between 21-29 years and over sixty percent, 46(63.6%) of patients were female. All the patients, 66(100%) had cough and chest pain, 57(86.4%) had crackles and about ten percent, 6(9.1%) had difficulty in breathing at the time of admission into the study. About 29(43.9%) of patients had fever at baseline and 14(21.2%) had a respiratory rate between 16-29 breaths per minute at baseline. At baseline, there were no significant differences between the two treatment groups in which the patients were allocated. (p >0.05). This is as summarized in Table 1

**Table 1:**
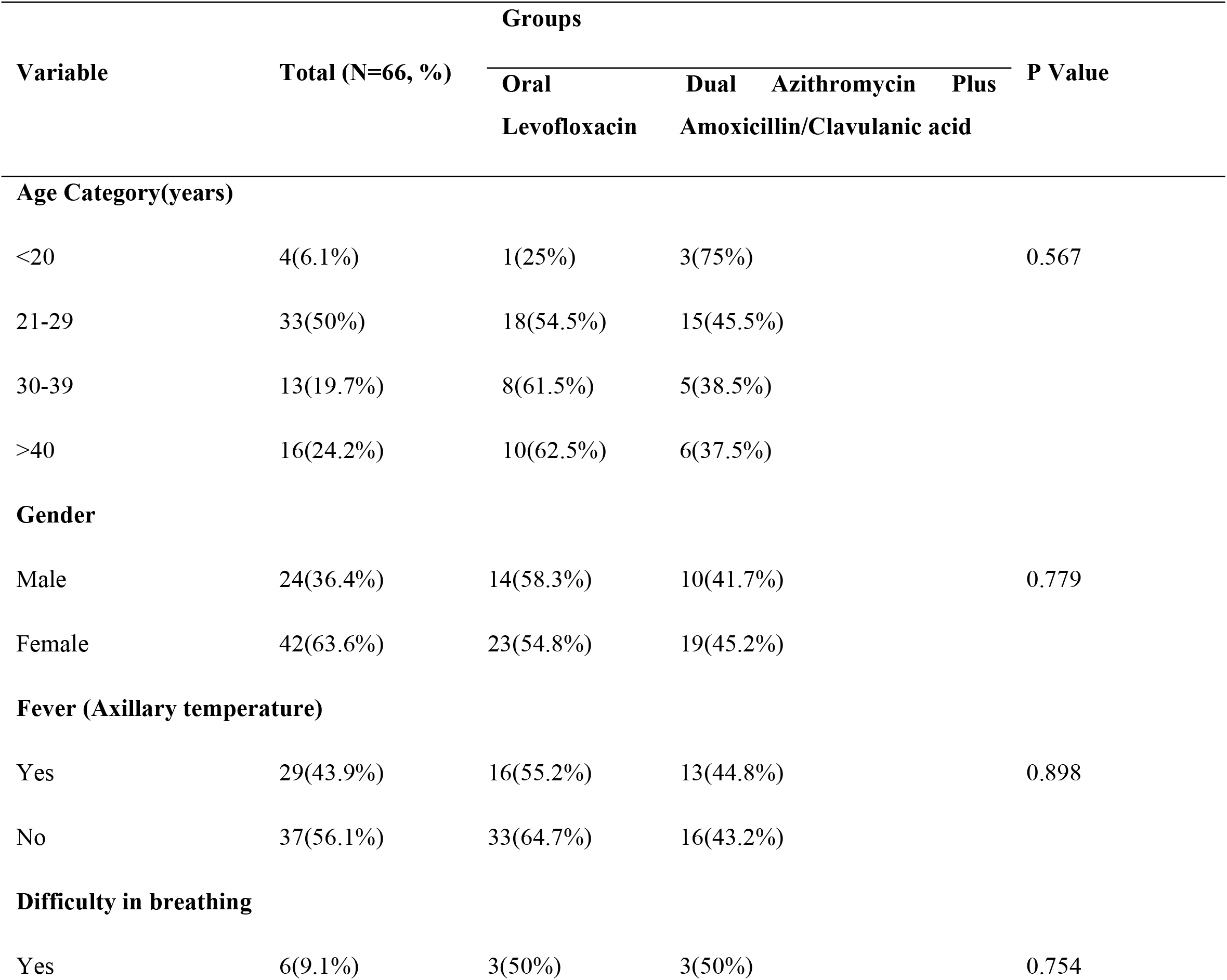
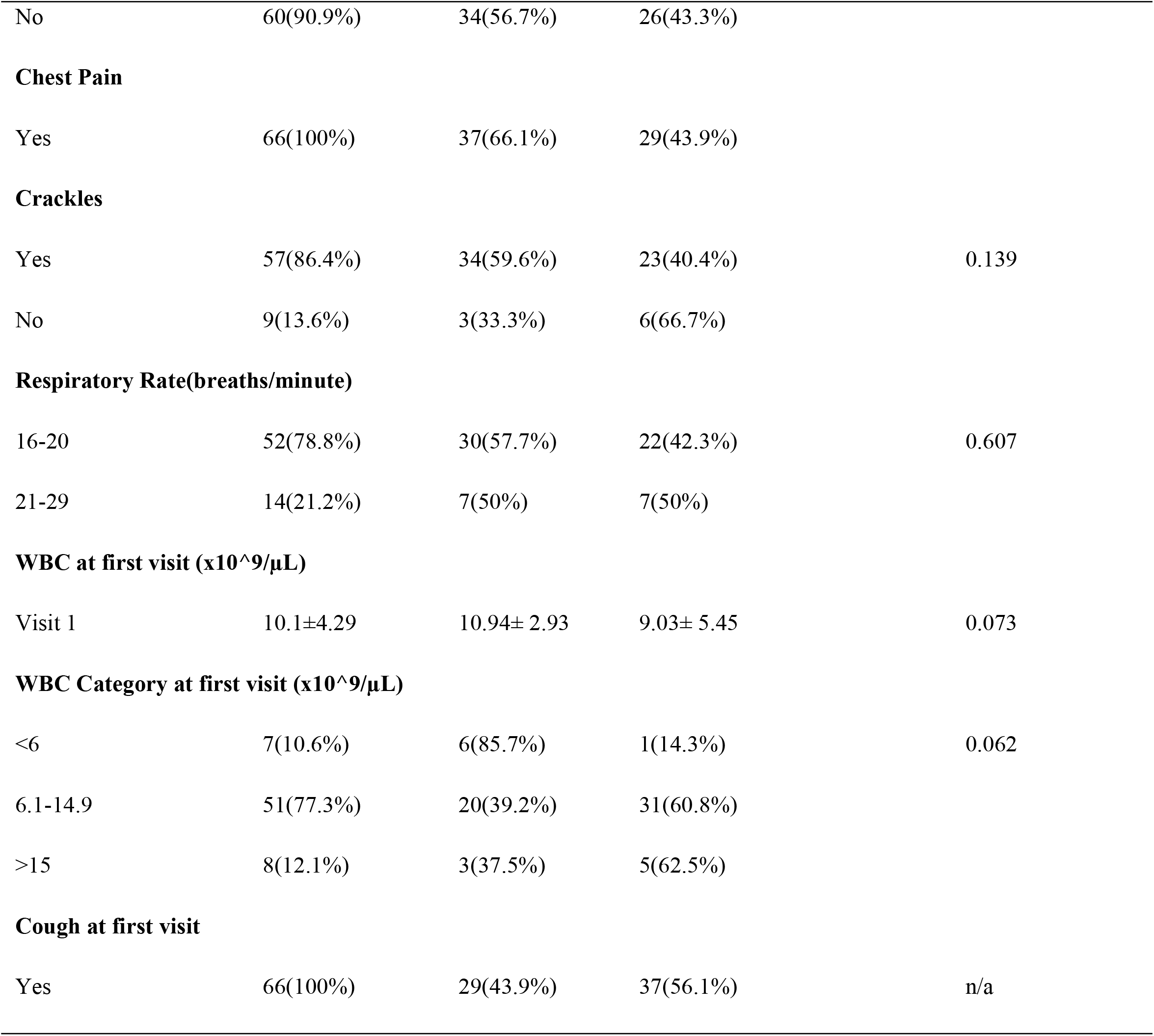
Demographic detail and Baseline Characteristics of the patients.

### Comparison of effectiveness of Oral Levofloxacin and dual Oral Azithromycin and Amoxicillin/Clavulanic Acid in treatment of CAP

#### Comparison of time to resolution of CAP as per WBC Count between the two treatment groups

At initial visit, about 8(12.1%) had elevated WBC count (>15x 10^9/µL) and about ten percent, 7(10.6%) had low WBC count (<6x 10^9/µL). At visit two and three, after start of treatment change in WBC count was significantly associated with the treatment group in which the patient belonged to. (p<0.05). At visit 2, 3(75%) of patients had elevated WBCs in the oral Levofloxacin group compared to only 1(25%) in the dual azithromycin amoxicillin/clavulanic acid group. At visit 3, one patient had elevated WBCs in the oral levofloxacin group whereas none had elevated WBCs in the dual Azithromycin and Amoxicillin/clavulanic acid group. This is as summarized in table 5

**Table 2:** WBC Count during the first visit and Subsequent visits.

| Variable | Total (N=66, %) | Groups |  | P Value |
| --- | --- | --- | --- | --- |
|  |  | Oral<br>Levofloxacin | Dual Azithromycin and<br>Amoxicillin/Clavulanic acid |  |
| WBC at Visit 1(x10^9/μL) |  |  |  |  |
| <6 | 7(10.6%) | 6(85.7%) | 1(14.3%) | 0.062 |
| 6.1-14.9 | 51(77.3%) | 20(39.2%) | 31(60.8%) |  |
| >15 | 8(12.1%) | 3(37.5%) | 5(62.5%) |  |
| WBC at Visit 2(x10^9/μL) |  |  |  |  |
| <6 | 12(18.2%) | 10(83.3%) | 2(16.7%) | 0.002 |
| 6.1-14.9 | 50(75.8%) | 16(32%) | 34(75.8%) |  |
| >15 | 4(6.1%) | 3(75%) | 1(25%) |  |
| WBC at Visit 3(x10^9/μL) |  |  |  |  |
| <6 | 19(28.8%) | 13(68.4%) | 6(31.6%) | 0.016 |
| 6.1-14.9 | 46(69.7%) | 15(32.6%) | 31(67.4%) |  |
| >15 | 1(1.5%) | 1(100%) | 0% |  |

#### Comparison of time to resolution of symptoms

There was a difference in mean time to resolution of cough and chest pain between the two treatment groups. The mean time to resolution of chest pain was 1.7 days in dual Azithromycin and amoxicillin /clavulanic acid group as compared to 2.21 days in oral Levofloxacin (p=0.009). The mean time to resolution of cough was 3.71 days in the oral Levofloxacin group as compared to 3.14 days in the dual Azithromycin and Amoxicillin/clavulanic acid(p=0.014). There was no difference in the meantime to fever resolution, time to crackles subsidence, time to resolution of difficulty in breathing, change in WBC count at visit 2 and 3 in oral Levofloxacin compared to dual Azithromycin and Amoxicillin/clavulanic acid (p>0.05). Table 6

**Table 3:** Comparison of effectiveness of oral Levofloxacin and the dual Azithromycin and Amoxicillin/Clavulanic acid based on time to resolution of symptoms.

| | Oral Levofloxacin<br>Group<br>(Mean±SD) | Dual Azithromycin and<br>Amoxicillin/Clavulanic<br>acid Group (Mean±SD) | Mean<br>Difference<br>(95% CI) | P value |
| --- | --- | --- | --- | --- |
| Time to Fever resolution(days) | 0.88±1.204 | 1.0±1.815 | -0.125(-1.216-0.966) | 0.817 |
| Time to Chest pain resolution(days) | 2.21±0.902 | 1.70±0.618 | 0.504(0.130-0.878) | **0.009** |
| Time to Crackles subsidence(days) | 1.00±0.853 | 1.11±0.583 | -0.114(-0.491-0.263) | 0.546 |
| Time to difficulty in Breathing resolution(days) | 0.10±0.310 | 0.11±0.393 | -0.005(-0.183-0.173) | 0.958 |
| Time to Cough resolution by day 5 | 3.71±0.588 | 3.14±0.789 | 0.568(0.123-1.013) | **0.014** |
| Change in WBC count at visit 2 | 7.53±4.072 | 11.18±13.31 | -3.654(-8.864-1.555) | 0.166 |
| Change in WBC count at visit 3 | 6.84±2.93 | 7.69±1.53 | -0.855(-1.973-0.261) | 0.131 |

## DISCUSSIONS

Out of the 16 (43.2%) of patients who had community acquired pneumonia and used azithromycin and amoxycillin/clavulanic acid, only 2 (5.4%) had fever at day five. At baseline, all patients in this group had chest pains 37 (100%) and none had chest pains at day 5. Crackles were resolved in all 34 (91.9%) patients at day 5. At the end of the treatment period (5-days) 9(24.3%) of the patients still had cough. These findings agree with (Caballero & Rello, 2011) who found that combination antibiotic therapy achieves a better outcome in the treatment of CAP. (Lodise et al., 2007) also found similar results that combining an extended-spectrum cephalosporin and a macrolide demonstrated better outcome in the treatment of community acquired pneumonia. Similarly, (Lee et al., 2018) found that combination therapy of the macrolides and beta lactam appears to result in improved survival and, possibly, shorter hospital length of stay in the hospital.

Levofloxacin appears to be a better choice in patients diagnosed with pneumonia and presenting with difficulty in breathing at first presentation. In this study, 10.3% of patients who had pneumonia with difficulty in breathing on the day of presenting to the hospital did not have difficulty in breathing on the subsequent days. Fever and crackles had resolved on the fourth day. However, cough and chest pains persisted in 41.4% and 10.3% of the patients respectively by the fifth day. This may mean that patients who have CAP who are treated with levofloxacin alone may need treatment period up to 7 – 10 days to achieve resolution of cough and chest pains. This study findings corroborate finding by (Wasserfallen et al., 2004) who found that the efficacy and tolerability of levofloxacin 500 mg once daily for 10 days in CAP patients is approved for use as monotherapy for CAP (Dunbar et al., 2004; Zar et al., 2020) had a similar findings with this study that levofloxacin therapy at 750mg once daily in community acquired pneumonia resulted in more rapid symptom resolution, with a significantly greater proportion of patients experiencing resolution of fever on the third day of therapy. Previous work, e.g. (Zar et al., 2020), explained that this is attributed to the fact that levofloxacin has better intracellular concentration. This also suggests that 500mg of levofloxacin twice daily has same efficacy as 750mg once daily.

The findings of this study suggest that both regimens used in the treatment of community acquired pneumonia demonstrated 100% cure rates. The differences were observed in symptoms such as cough, chest pains and fever. Dual azithromycin and amoxycillin/clavulanic acid showed superior results to levofloxacin in resolution of cough and chest pains at 75.6% vs 58.6% and 100% vs 89.6% respectively. Levofloxacin demonstrated better outcomes in fever resolution at the end of the 5 days (p=0.817). There was significant change in white blood cell count from baseline at the end of the treatment in all the treatment groups (p=0.062) at baseline versus (p=0.016) on day 5 of treatment with levofloxacin group having one patient demonstrating elevated WBC compared to none in amoxycillin/clavulanic acid combined with azithromycin. The findings of the current study showed that amoxycillin/clavulanic acid combined with azithromycin showed a modest superiority to levofloxacin in the treatment of community acquired pneumonia although not statistically significant across rate of symptom resolution and change in white blood cell count.

## CONCLUSIONS

Although all the drugs showed better results when used to treat community acquired pneumonia, azithromycin combined with amoxycillin/clavulanic acid demonstrated superior results compared to levofloxacin alone.

## RECOMMENDATIONS

Based on these conclusions, the practitioners and policy makers should;

i. Prioritize the use amoxycillin/clavulanic acid combined with azithromycin in treatment of community acquired pneumonia in patients who have had previous antibiotic exposure within the last three month.
ii. Not restrict the use of levofloxacin in patients who may benefit in treatment of community acquired pneumonia provided the possibility of tuberculosis has been ruled out. This is because levofloxacin alone has demonstrated almost similar efficacy to amoxycillin/clavulanic acid and azithromycin.

To better understand the implications of the results in this study, future studies could address the optimal dosage for patients using levofloxacin (500mg twice daily versus 750mg daily).

## Data Availability

All relevant data are within the manuscript and its Supporting Information files.

NA

